# Multiple cohort study of hospitalized SARS-CoV-2 in-host infection dynamics: parameter estimates, sensitivity and the eclipse phase profile

**DOI:** 10.1101/2022.06.20.22276662

**Authors:** Chapin S. Korosec, Matthew I. Betti, David W. Dick, Hsu Kiang Ooi, Iain R. Moyles, Lindi M. Wahl, Jane M. Heffernan

## Abstract

Within-host SARS-CoV-2 modelling studies have been published throughout the COVID-19 pandemic. These studies contain highly variable numbers of individuals and capture varying timescales of pathogen dynamics; some studies capture the time of disease onset, the peak viral load and subsequent heterogeneity in clearance dynamics across individuals, while others capture late-time post-peak dynamics. In this study, we curate multiple previously published SARS-CoV-2 viral load data sets, fit these data with a consistent modelling approach, and estimate the variability of in-host parameters including the basic reproduction number, ***R***_**0**_. We find that fitted dynamics can be highly variable across data sets, and highly variable within data sets, particularly when key components of the dynamic trajectories (e.g. peak viral load) are not represented in the data. Further, we investigated the role of the eclipse phase time distribution in fitting SARS-CoV-2 viral load data. By varying the shape parameter of an Erlang distribution, we demonstrate that models with either no eclipse phase, or with an exponentially-distributed eclipse phase, offer significantly worse fits to these data, whereas models with less dispersion around the mean eclipse time (shape parameter two or more) offered the best fits to the available data.

## 1 Introduction

Since late 2019, the SARS-CoV-2 virus has significantly disrupted life globally [1–3]. Two years later, many countries are moving to consider SARS-CoV-2 and the resulting COVID-19 infection to be endemic [4], which will require new strategies for forecasting, management and control. An endemic state will necessitate a sustained reliance on data fitting [4], and on analytical tools, as public health authorities manage risk and allocate resources in future years.

Mathematical models have been employed in the estimation of key parameters of the evolution of SARS-CoV-2 outbreaks at the population level. Mathematical models are able to predict key parameters like the time dependent epidemiological reproduction number [5], the effects of quarantine and risk of importation [6], the effects of aggregate non-pharmaceutical intervention [5], the risks associated with relaxing interventions competing with increasing vaccination [7], and the effects of vaccination waning and boosting on case loads [8]. All of these estimates can then be used with mathematical models that estimate the impact of COVID-19 on healthcare systems [9].

Mathematical modelling has also been used to model SARS-CoV-2 in-host infection dynamics. Mathematical models at the in-host level can estimate parameters and outcomes that may be difficult to measure at the population level. For example, SARS-CoV-2 in-host models have been used to model the immune response generated by vaccines, to predict which populations vaccines are likely to impact most [10], to estimate the efficacy of vaccines in individuals [11], as well as to model T cell dynamics and cytokine secretion in mild, moderate and severe cases of COVID-19 [12]. A major focus of SARS-CoV-2 in-host modelling has been understanding viral load dynamics: examples include correlating viral load with mortality [13]; linking viral load dynamics with host transmission [14, 15]; understanding the effects of antiviral therapy on the shedding dynamics [16]; assessing individual-level heterogeneity in mild cases [17]; and relating viral load with viral replication and the immune response [18].

Recent studies have found that viral load levels in the upper respiratory track are similar in symptomatic and asymptomatic individuals [19], and further, that viral load can vary by five orders of magnitude and yet still display no correlation with disease severity [18]. In contrast, others have reported that high viral load titres are correlated with an increase in mortality as well as disease severity [20–24], suggesting that case severity (hospitalized versus non-hospitalized) may be an important factor in viral load. Furthermore, the time to peak, as well as magnitude of peak viral load, are informative in understanding the probability of transmission and when transmission is likely to occur throughout the course of the disease [20, 25, 26]. Correctly characterizing viral load dynamics with an accurate mathematical model is critical in accurately predicting disease outcomes and transmission dynamics.

It is important to note that SARS-CoV-2 in-host viral load studies can include cohorts of various sizes, in which individual shedding dynamics are collected at various points in time throughout the disease time-course and across the pandemic, both temporally and geographically. In addition, due to difficulties inherent in early data collection, many studies capture limited dynamics of the trajectory of viral load within a single individual. Using mathematical modelling and statistical fitting methods, in-host models and parameter estimation, relationships between different viral load data sets can be uncovered.

In the current study, we curated three previously published viral load data sets for which temporal viral load trajectories were available [13, 16, 27], and fit each data set to a series of target-cell limited in-host models. The data sets used consist of primarily hospitalized individuals, and vary in size from 25 to 4344 individuals. Fitting these data sets to the same set of in-host models allows us to quantify the reliability and reproducibility of fitted parameter values, both within and among studies. We are also able to address how real-world constraints such as data sets containing fewer individuals, or data sets in which the peak response was not captured, affect the uncertainty of the fitted parameters and estimated reproduction number, *R*_0_.

Previous SARS-CoV-2 in-host studies have employed an exponentially-distributed eclipse phase to describe viral load dynamics [13, 15, 16, 28–30]. The eclipse phase is included to model the delay between successful target cell infection and the production of virus; biologically, the input virion is “eclipsed” by the cell, initiating a multitude of processes that ends when the cell is able to productively bud new virions. The mean duration of this process, referred to as the mean eclipse time, varies between diseases, cell types, and most likely across various strains of the same virus. For example, the length of the eclipse phase for influenza virus has been found to vary between 6 to 12hr depending on the strain [31–34]. The probability distribution for duration of the eclipse phase takes into account the multiple timescales for the succession of biological mechanisms that end in viral budding, and also accounts for the stochastic variability inherent to these processes [35]. Incorporating a single eclipse compartment, as done in previous SARS-CoV-2 viral load studies [13, 15, 16, 28–30], implicitly assumes an exponentially-distributed eclipse duration. Biologically, this corresponds to the unrealistic situation in which some cells will instantaneously enter a budding state at the moment of infection, neglecting eclipse dynamics entirely. The probability distribution for the eclipse phase duration for SHIV, for example, has been found to best be described by a fat-tailed distribution [36], and for influenza by a normal or log-normal distribution [32]. Therefore, another focus of this work was to fit multiple published SARS-CoV-2 data sets to models with varying eclipse time distributions to assess via log-likelihood estimators (including penalties for additional parameters) if a particular model is preferred.

## 2 Methods

### 2.1 Clinical data acquisition and summary

All viral load data sets used in this work were previously published, the details of which are summarized in Table 1. If unavailable directly from the published source, we digitized the data using the software WebPlotDigitizer (version 4.5) [37]. All data sets are in units of log_10_ copies per mL. For all data sets we fit to cohorts of individuals, that is, viral load measures were grouped and fit separately for each study, but not for each individual within a study.

**Table 1.**
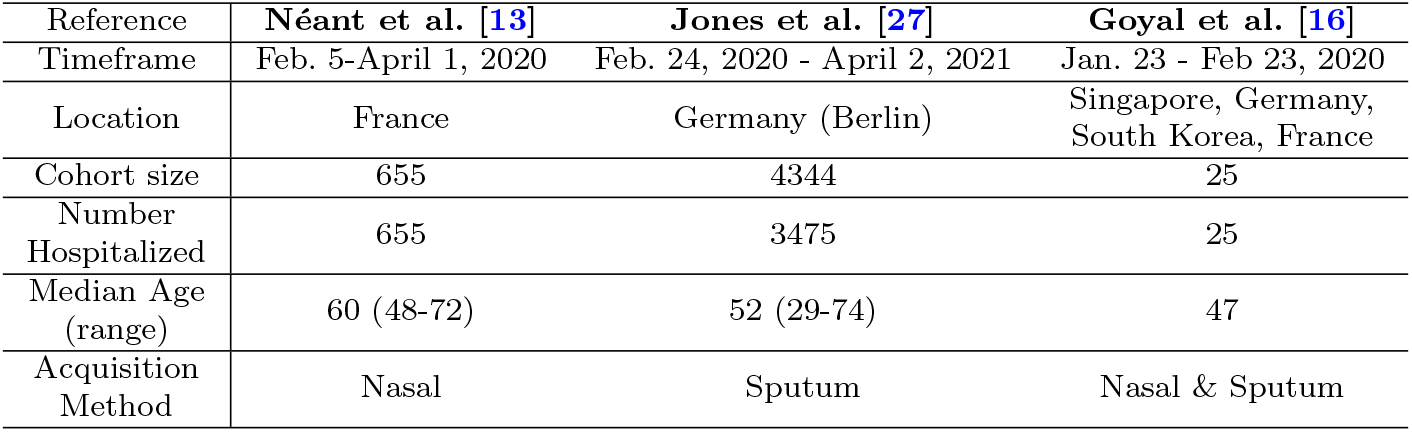
Summary of individual SARS-CoV-2 viral load cohorts used in this work.

The Goyal data set [16], consisting of 25 individuals whose viral load time courses were collected from a mix of throat and nasopharyngeal swabs, was obtained from various sources: 11 hospitalized individuals from Singapore, median age 47 (31-73), symptom onset ranging from January 14-30, 2020 [38]; 9 hospitalized individuals from Munich, Germany, described as young to middle aged, symptom onset occurring in January, 2020 [39]; 1 hospitalized individual from South Korea, aged 35 years old, symptom onset 18th of January, 2020 [40]; 4 hospitalized individuals from Paris, France, median age 46, symptom onset mid-January, 2020 [41]. Measurement times in this data set correspond to days after first positive test. For the fits to this data set completed in this work, all values below the limit of 2 log_10_ copies/mL were considered below the detection threshold and censored.

The Néant data set [13] includes data obtained from 655 hospitalized individuals with a median age of 60. Viral load measurements were collected via nasal swabs from February 5 to April 1 2020 in France, where measurement times correspond to days since symptom onset.

The Jones data set [27] consists of viral load measurements from 4344 individuals, 3475 of which were hospitalized, with a median age of 52. Viral load measurements were acquired by sputum swabs from February 24, 2020, to April 2, 2021 in Berlin, Germany. Measurement times correspond to days since peak viral load. We note that Jones *et al* [27] report on a much larger data set consisting of 25,381 individuals, but for this work we only considered the subset of those (*n* = 4344) for which viral load time courses were available. Jones *et al*. [27] report that 80% of the 4344 were hospitalized.

### 2.2 Target-cell limited model

To model the within-host dynamics of SARS-CoV-2 viral shedding we use a target-cell limited model similar to that previously used to model influenza A [42] and SARS-CoV-2 [13, 28]. The model includes target cells (*y*_*t*_), productively infected cells (*y*_*B*_) capable of budding infectious virus (*v*) and non-infectious virus (*w*), and non-productively infected cells in the eclipse state, which is extended to include *k* eclipse stages (*y*_*k*_). Target cells are assumed to be infected by infectious virus at rate *α* (mL· d^*−*1^ · copies^*−*1^). Newly infected cells enter the eclipse phase. We consider a range of eclipse time distributions, each of which has a mean duration that is fixed to 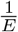 days. In particular, we extend the typical one-compartment model, which yields exponentially-distributed eclipse durations, to a linear chain of *k* eclipse stages [43], yielding Erlang-distributed eclipse durations with shape parameter *k*. At the end of the eclipse phase, infected target cells enter the productively infected cell class, *y*_*B*_. These cells have a constant loss rate *D* (*d*^*−*1^) and produce virions at rate *B* 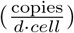. Following the SARS-CoV-2 modelling work of Néant *et al*. [13], we assume that a fraction, *E*, of virions are infectious, and the remaining (1-*E*) are non-infectious. Both infectious and non-infectious virions are cleared at a constant rate of *C* (*d*^*−*1^). A schematic of this target-cell limited model is shown in Fig. 1a, and is given by the following system of ordinary differential equations:

**Fig. 1.**
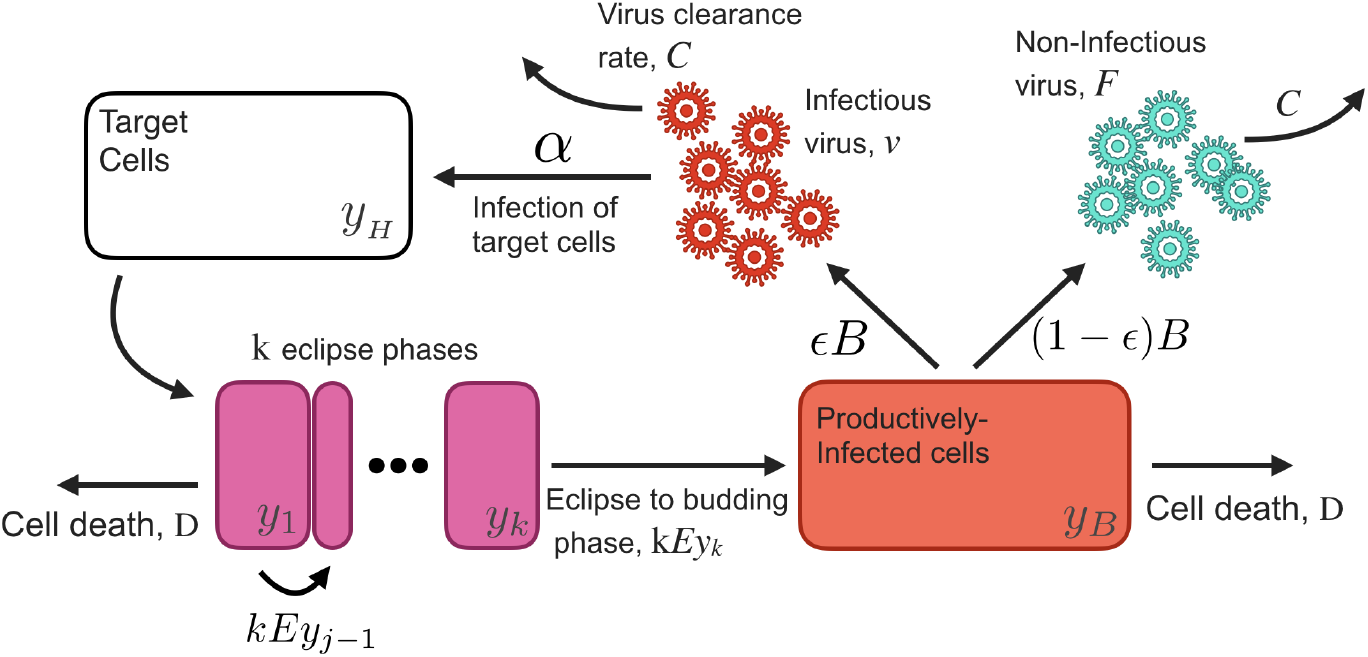
Schematic of target-cell limited model described in section 2.2.

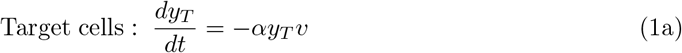

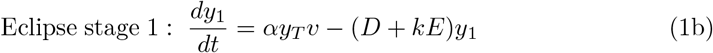

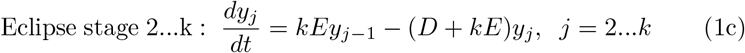

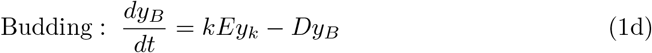

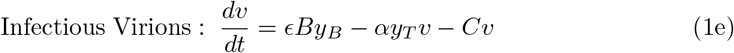

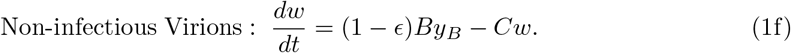

We also consider a reduced model with no eclipse phase; in this case newly infected cells instantaneously become productively infected (budding) cells. The no-eclipse model is:

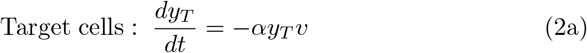

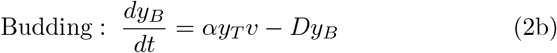

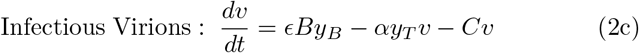

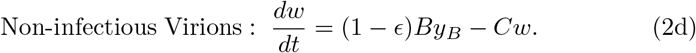

### 2.3 Assumptions on initial conditions and parameter values

For each cohort fit, we assume an initial time of infection *t*_inf_ (days), where *t*_inf_ ≤ 0, and where *t*_inf_ is determined through fitting with bounds on the fit of 0-14 days. Thus *t*_inf_ represents an expected initial time of infection for the cohort and the standard error in the estimate of this parameter may reflect heterogeneity in initial times across individuals. We assume an initial target cell concentration of 1.33×10^5^ cells mL^*−*1^ as used in previous studies [13, 28, 44]. This value is obtained by assuming the upper respiratory tract contains 4×10^8^ cells distributed evenly in a volume of 30 mL, and where only 1% of the cells express the ACE2 receptor associated with viral entry [45]. We assume an initial condition of one productively infected cell in the upper respiratory tract (*y*_*B*_ = 1 cell/30 mL). The initial condition for all *y*_*j*_ eclipse states is 0. The initial number of infectious and non-infectious virus particles is assumed to be 0.

We assume the proportion of produced infectious virions to be fixed at *ϵ* = 10^*−*4^, which was determined to be the upper bound in SARS-CoV-2 infection in non-human primates [29]; non-infectious virions are therefore produced at rate (1−*ϵ*). SARS-CoV-2 viral titres have been measured as soon as 2 hours post infection [46]; as not all parameters in the model can be fit, we fix the mean eclipse duration to 1*/E* = 0.2 days, which corresponds to viral production beginning, on average, 4.8 hours after cell infection. For each cohort we fit the parameters *α*, B, C, and D. A summary of all parameter assumptions and initial conditions can be found in Table S1.

### 2.4 Parameter estimation and fitting assessment

All fits to Eq. 1 were performed in Monolix [47] (Version 2020R1) using non-linear mixed-effects models. Individual parameters for each data set are determined by the maximum likelihood estimator Stochastic Approximation Expectation–Maximization (SAEM), and all fits met the standard convergence criteria (complete likelihood estimator). For all data sets we fit the parameters *α, B, C, D* and *t*_inf_, and all parameters were assumed to be lognormally distributed. Further details of the fits, including priors, population values, and random effects, are given in Table S1.

### 2.5 Sensitivity and error analysis

We report residual errors assuming a ‘combined’ error model, whereby the error term in the observation model is composed of a constant fitted term and a term proportional to the structural model [47].

We perform sensitivity analysis to characterize the response of model outputs to variation in the fitted parameters as well as the fixed parameter, *E*. Latin hypercube sampling (LHC) and Partial Rank Correlation Coefficient (PRCC) [48] are employed to study the effects of model outcomes on the peak value of each state variable.

An independent error analysis is also completed to complement the fit errors found by Monolix. Here we compute the mean standard error between the curve produced by the target-cell limited models and the Jones data set [27]. For each set of fitted parameters, for each value of *k*, the mean standard error is computed by

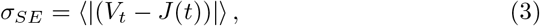

where *V*_*t*_ = log [*v*(*t*) + *w*(*t*)] and *J* (*t*) is the average value from the Jones data set at time *t*. To compute *σ*_*SE*_, we bin the Jones data in 0.5 day intervals, where the data in each interval is averaged.

## 3 Results

### 3.1 Cohort-dependent variation in viral load dynamics

A schematic illustrating our model for *k >* 0 is shown in Figure 1. Figure. 2a depicts the timeline of the studies used in this work, which range from January 2020, to April 2021, and also vary both geographically and in cohort size (details provided in Table 1).

**Fig. 2.**
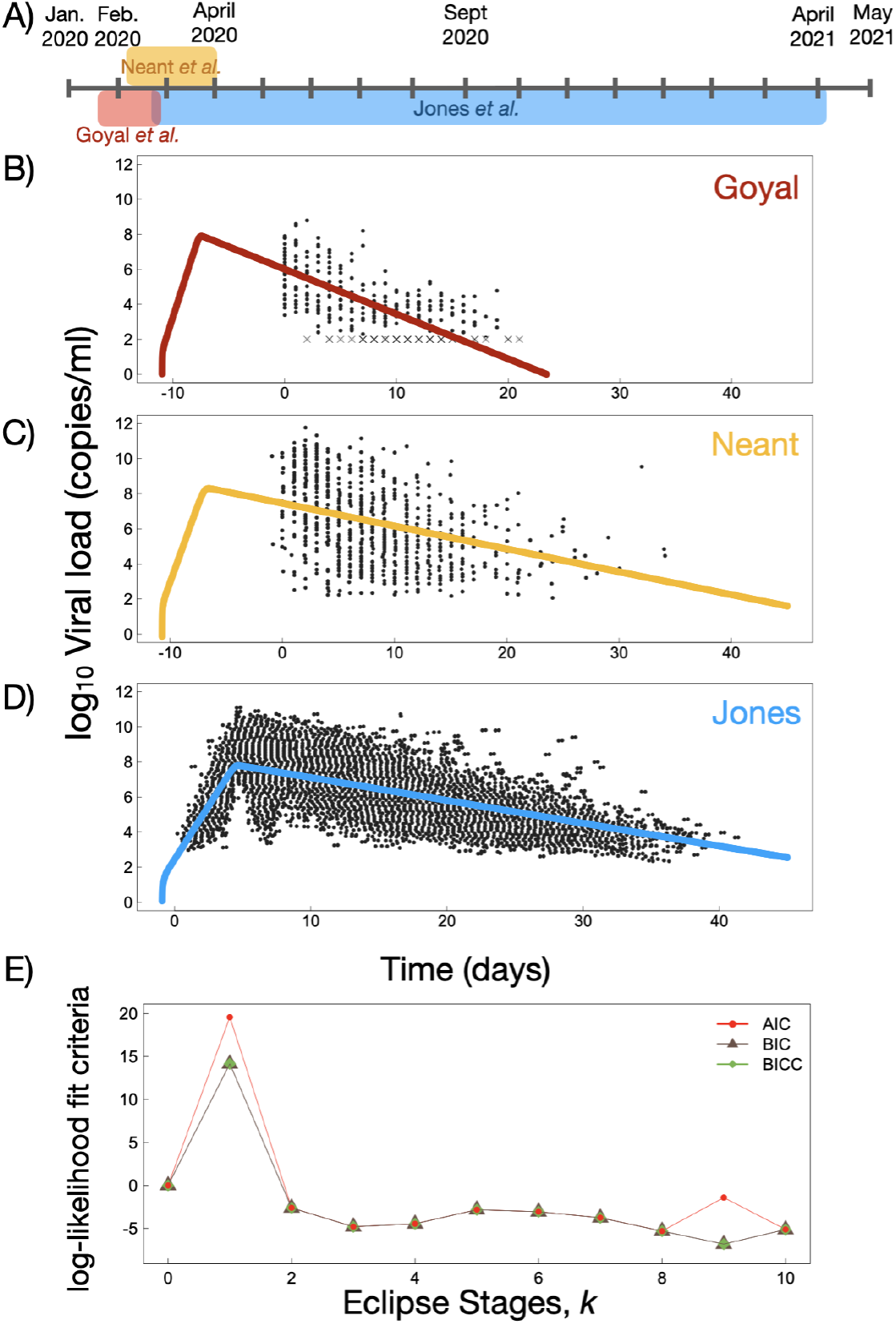
(A) Timeline of viral load studies used in this work. Further details about each study can be found in Table 1. (B)-(D) log_10_ viral load and examples of cohort fits for each data set used in this work. (E) Log-likelihood criteria for best fit as a function of number of eclipse stages, *k*. The value of each criterion at *k* = 0 has been subtracted for visual comparison.

Examples of fits to all data sets are shown in Fig. 2b-d. In Fig. 2e we report, as a function of *k*, three different log-likelihood fit criteria: AIC (Akaike information criterion), BIC (Bayesian information criterion), and BICC (corrected Bayesian information criterion). Noting that a lower log-likelihood criterion suggests a more preferred model, we find non-monotonic behaviour in all criteria as a function of increasing *k*. The BIC, BICC, and AIC all increase from *k* = 0 to *k* = 1, and then decrease to a minimal plateau from *k* = 2 to *k* = 8. We find that from *k* = 8 to 10 the BIC and BICC values are approximately constant, while the AIC displays a slight increase at *k* = 9. Irrespective of the log-likelihood criterion used, the best fit occurs when *k* = 3. However values of all the log-likelihood parameters are within ≈ 2 units of the minimum log-likelihood for *k* = 2 through *k* = 8, suggesting that models with two through eight eclipse stages cannot be rejected as being the most appropriate model to fit these data (see Discussion for further details).

In Fig. 3 we plot the best fit parameter values as a function of *k* for the Goyal, Jones and Néant cohorts, while Fig. S1 displays the estimated time of infection, *t*_inf_. The error bars in Fig. 3 show the residual error for each parameter determined by the fit [47], and open symbols give the mean and standard deviation of best fit parameters across all accepted models (*k* = 2…8). The best fit values of each parameter for each data set, as well as the mean and standard deviation across all *k* values, are provided in Table S2.

**Fig. 3.**
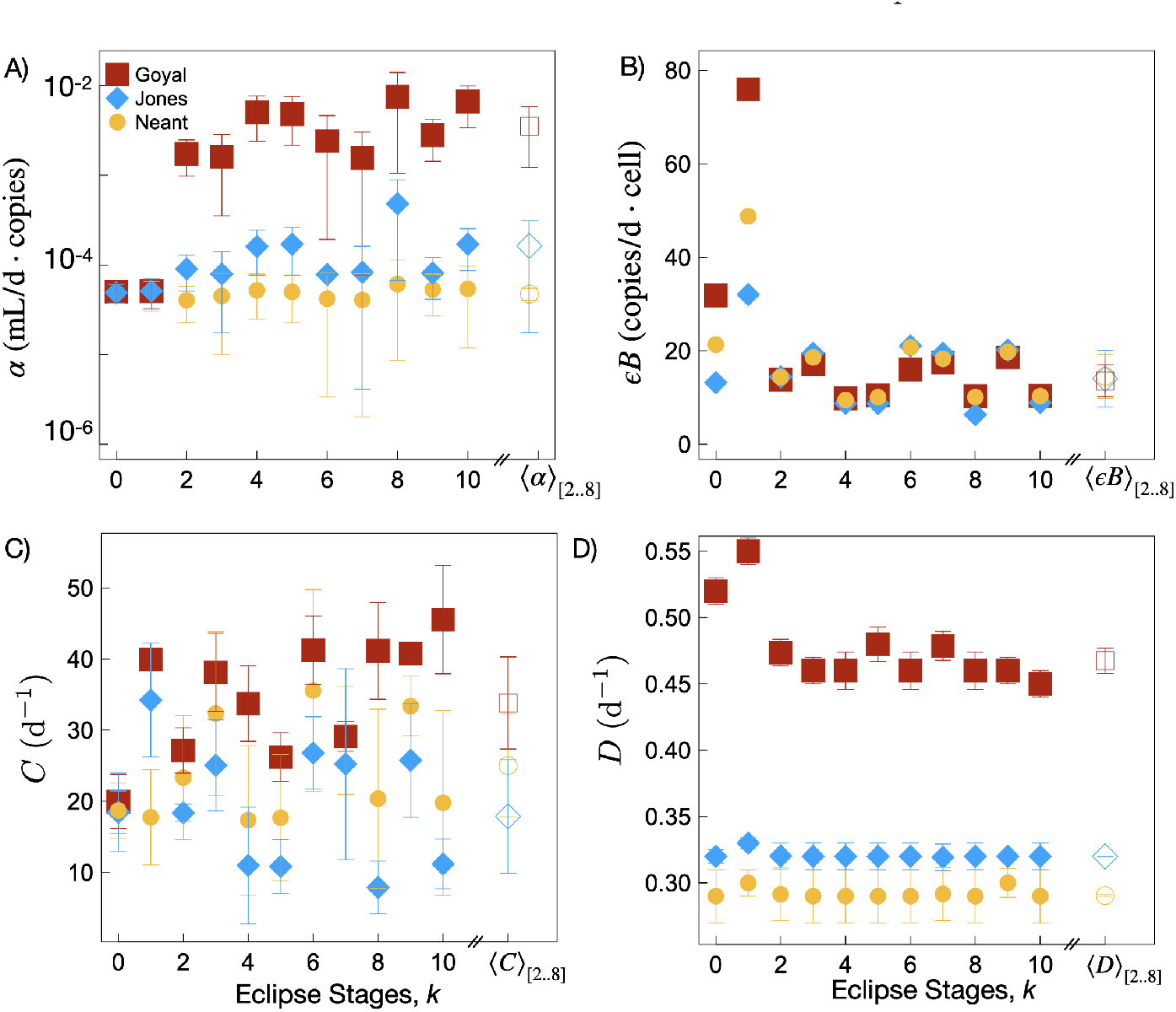
Best-fit parameter values for the Goyal, Néant and Jones data sets as a function of the number of eclipse stages, *k*. Error bars are the residual error determined through the fit. Open symbols (on the right) of each plot are the mean and standard deviation for each parameter value across all accepted fits (*k* = 2…8). (A) The per-target cell attachment rate, *α*. (B) The infectious virion budding rate, *εB*. (C) The virion clearance rate, *C*, and (D) The cell removal rate, *D*.

A general conclusion from Fig. 3, consistent with the results from the log-likelihood criteria, is that best-fit parameter values for *k* = 0 and *k* = 1 can vary substantially from best-fit values obtained for *k* ≥ 2. Focusing on the results from accepted models (open symbols) we see that the estimated budding rate, *εB*, is fairly similar across cohorts, and is relatively insensitive to *k* for *k* ≥ 2. The best-fit values of *α* and *D* are also relatively robust to changes in *k* for *k* ≥ 2, but vary substantially across the different cohorts. Finally, the parameter *C* shows substantial variability among cohorts, and within each cohort shows a high sensitivity to *k*.

A Partial Rank Correlation Coefficient sensitivity analysis is performed to assess how model fit parameters affect the peak response from each state variable, the result of which is shown in Figure S5. We find the sensitivity analysis to reveal intuitive trends based on the model structure for all fitted parameters. LHS parameters close to +1/-1 indicate strong influence on the outcome measure, where negative values suggest the parameter is inversely proportional to the outcome measure, and PRCC values whose magnitude is greater than 0.5 are considered important [49]. We find peak *v* and *w* have positive PRCC values of ∼1 for the budding rate, *B*, and less than − 0.5 PRCC values for the clearance and cell removal rates *C* and *D*, respectively. We also find the *y*_*B*_ peak to have a PRCC value of less than − 0.5 for the cell removal rate, *D*.

### 3.2 *R*_0_ analysis

The basic reproduction number, *R*_0_, is defined by the average number of secondary infected cells resulting from a single infected cell in a population of susceptible cells at the beginning of infection [50–52]. Following the method of Diekmann and Heesterbeek [50, 51], we derive *R*_0_ for our target-cell limited model (Eq. 1) for *k* eclipse phases (*k*≥1) assuming initial disease-free equilibrium conditions (see supplementary material for calculations of the disease-free equilibrium). For *R*_0_ corresponding to Eqs. 1 we use the notation 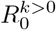. We find 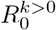 to be

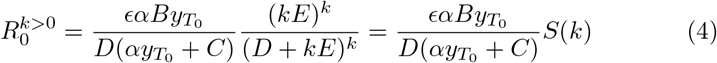

where *S*(*k*) denotes the probability that an infected cell survives all *k* eclipse stages, since the probability of maturing through a single eclipse stage rather than dying is given by *kE/*(*kE* + *D*). In the limit of *k* = 0, corresponding to

Eqs. 2,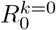 is found to be

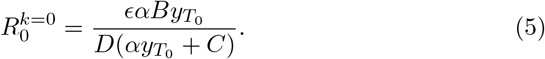

Thus, 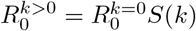. Therefore, the effect of the eclipse phase on the basic reproduction number is simply to reduce *R*_0_ by S(k).

Fig. 4a displays 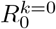 and 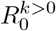 as a function of the number of eclipse stages, *k*, for all three data sets. We find the Néant and Jones data sets to have estimated *R*_0_ values of ∼10 and ∼17.5 across all accepted model fits (*k* = 2…8). As reflected in the best-fit parameter estimates, the use of *k* = 0 or *k* = 1 can result in markedly different predictions for *R*_0_; this is particularly striking in the Néant data set. The *R*_0_ estimate obtained for the Goyal data set shows substantial sensitivity to the choice of *k*, which may be a result of the small cohort size, and limited number of data for this study.

**Fig. 4.**
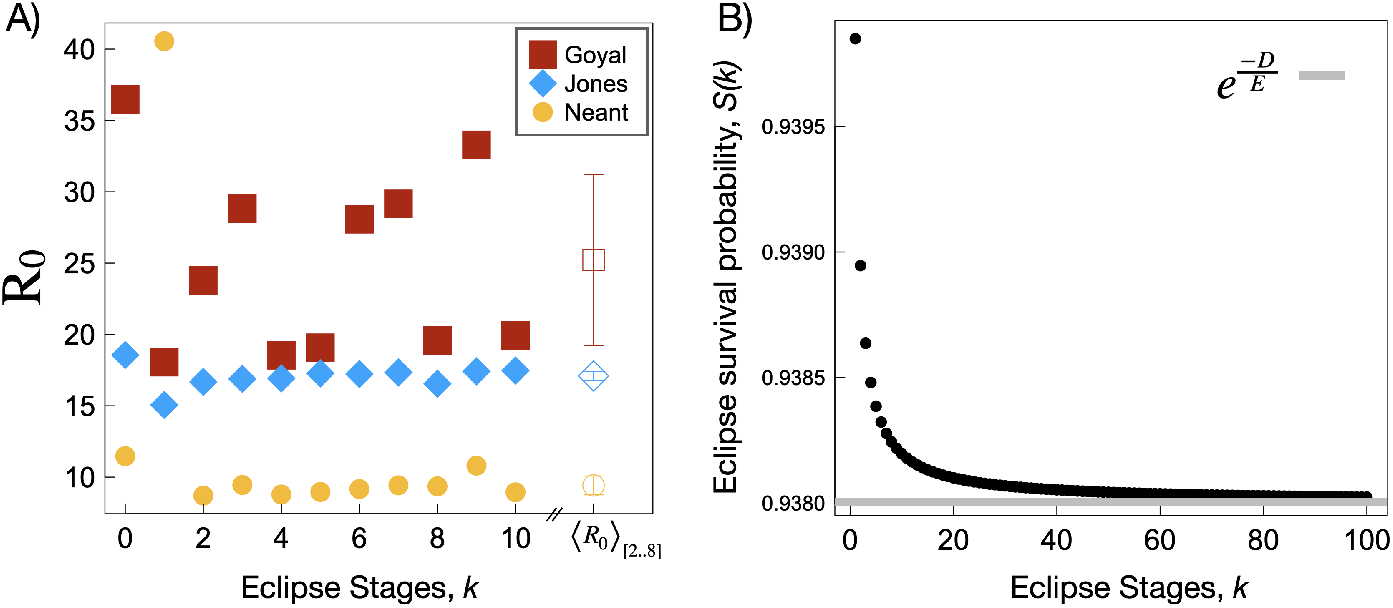
(A) *R*_0_ as a function of k for the best-fit parameter values for each data set. Open symbols (on the right) show the mean and standard deviation of *R*_0_ across all accepted fits (*k* = 2…8). (B) The probability of surviving the eclipse phase, *S*(*k*) versus the number of stages in the eclipse phase, *k. S*(*k*) approaches the limit 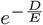 as *k → ∞*.

To gain further insight into the influence of the fitted parameters *α, B, C*, and *D* on *R*_0_ with increasing *k*, we compute 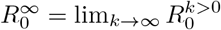. Using a straightforward application of L’Hôpital’s rule, we obtain

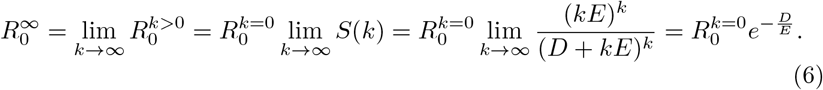

Fig. 4b shows the approach of *S*(*k*) to the limiting value of 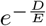 given parameter values *D* and *E* of 0.32 and 5 d^*−*1^, respectively. This figure shows that as the number of eclipse stages increases, *S*(*k*) monotonically decreases towards the 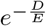 limit.

We point to the narrow *y*-axis scale in Fig. 4b, showing that, for the parameter values illustrated here, varying the number of eclipse stages over a large range has a negligible effect on *R*_0_. In particular, fixing *E* = 5 d^*−*1^, and taking *D* = 0.29, 0.32, or 0.48 as estimated for the Néant, Jones and Goyal studies respectively (Table S2), changing *k* from 0 to the limit as *k* → ∞, we find a maximum percent reduction in *R*_0_ of 5.3, 5.8 and 8.3% in these three studies, respectively.

Although *R*_0_ is relatively insensitive to *k* at fixed parameter values, we also note that as *k* changes, the best-fit parameter values may also change. Nonetheless as seen in Fig. 4a, estimates for *R*_0_ are relatively insensitive to *k* for the Jones and Néant data sets across the accepted models (*k* = 2…8).

In Fig. 5 we plot 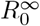 as a function of *α, ϵB, C*, and *D*. Coloured regions highlight the standard deviation of parameter estimates across all models for best-fit parameters for each of the three data sets, while the solid coloured vertical lines are the mean parameter estimates. The steepness of the curve in each region thus reflects the sensitivity of *R*_0_ to changes in each parameter. For example we find that the best-fit *α* values from the Goyal cohort lie in a region of parameter space where changes in *α* have little effect on *R*_0_, whereas slight changes in the *α* value estimated for the Néant cohort may appreciably affect *R*_0_ (Fig. 5a). Clearly 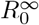 (Eq. 6) has a linear dependence on *B* with constant slope. Thus, variations in *B* for any study would lead to a similar shift in *R*_0_ (Fig. 4b). 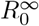 monotonically decreases as a function of both *C* and *D*; the Goyal fits are found to have the highest values for both parameters, whereas the Néant and Jones fits have quite similar values for all *k*. Thus, this analysis suggests that the larger values of *R*_0_ estimated for the Goyal cohort may be largely attributed to the larger *α* values estimated for that data set, as compared to the Néant and Jones data sets (Fig. 3a).

**Fig. 5.**
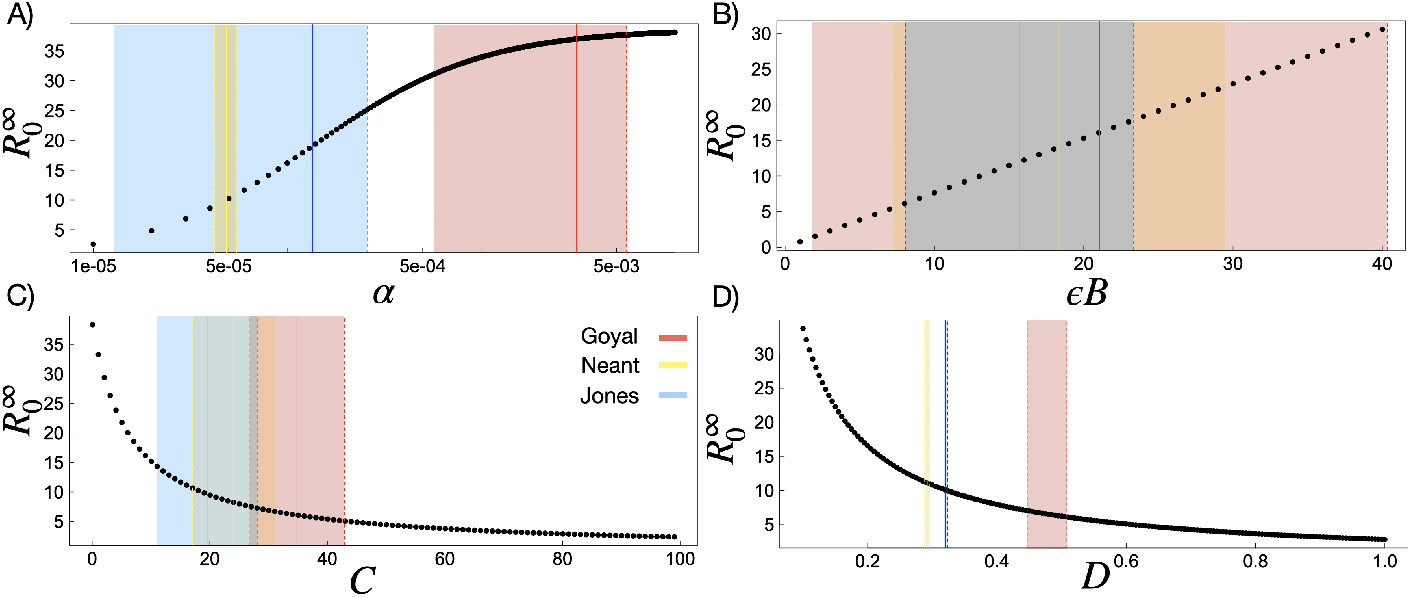
(A-D) 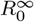 (Eq.6) as a function of *α, εB, C*, and *D*, respectively. Coloured regions corresponding to the standard deviation of parameter estimates across all models, while the solid vertical lines are the mean parameter values across all models. For best fits to the Goyal, Néant, and Jones data sets for all *k >* 0 are shown for illustrative comparison.

## 4 Discussion

We employed a series of target-cell limited models accounting for infected cell budding, a loss rate of infected cells, differing eclipse stage dynamics, and infectious and non-infectious virion production and clearance. We fit these models to three previously published SARS-CoV-2 viral load cohorts comprised of mostly hospitalized individuals: Néant [13] (n = 655), Jones [27] (n = 4344) and Goyal [16] (n = 25). We varied the number of eclipse stages, *k*, from 0 (Eqs. 2) to 1-10 (Eqs. 1), where all fits consist of the same number of fitted parameters, and begin with similar priors (See Table S1 for this information). All fits were completed in Monolix [47] which employs the SAEM algorithm, and all fits met the log-likelihood convergence criteria (see Methods for details).

Eclipse dynamics capture the inherent delay between the moment that a virion infects a target cell and the time at which the cell begins to bud new virions. A single eclipse compartment (*k* = 1) yields an exponentially-distributed eclipse duration, and is often implemented in in-host models as the simplest way to incorporate an eclipse stage (see [53]). This “simplest approach” to model development is an important guiding principle, particularly when faced with noisy, real-world clinical data. Increasing the number of freely fit parameters can lead to overfitting and may potentially introduce identifiability issues; interpreting the biological consequences in the resulting parameter fits becomes more difficult or impossible. Although extending the model from *k* = 0 to *k* = 1 involves introducing a new parameter – the eclipse rate *E* – we fixed this parameter based on previous SARS-CoV-2 literature [46] for all *k >* 0 to allow for a more fair comparison between models for *k* = 0 and *k >* 0. Thus, we fit the same number of parameters, each with consistent priors, for all models.

While the lowest AIC obtained among a group of candidate models (AIC_min_) indicates the most preferred model, the amount by which the AIC of a particular model exceeds this minimum (ΔAIC = AIC - AIC_min_) is a key parameter in accepting or rejecting candidate models. A value of ΔAIC of less than 2 reflects ‘equal support’ for the two models. ΔAIC of 2-10 is considered ‘substantial support’ for the less preferred model, while ΔAIC of 10 or more reflects ‘essentially no support’ [54]. For our models, all three log-likelihood criteria increase substantially from *k* = 0 to *k* = 1, followed by a decrease to a near-equivalent preference for *k* ≥2. We find that *k* = 3 is the most preferred model with the lowest AIC (Fig. 2e). Values of ΔAIC for *k* = 2…10 (excluding 3) vary between 0.4 (*k* = 4) and 3.4 (*k* = 9). Thus, although *k* = 3 results in the lowest AIC, all *k >* 1 models considered in this study are either considered ‘equivalently preferred’ or have ‘substantial support’ as an acceptable model. For *k* = 0, ΔAIC = 4.8, suggesting that the inclusion of an eclipse phase is justified in fitting these data. However for *k* = 1, ΔAIC =24.4; thus the use of a single eclipse compartment (exponentially-distributed eclipse duration) can be strongly rejected.

The finding that models containing *k >* 1 eclipse stages are preferred over *k* = 1 suggests that the eclipse duration is not best-described by an exponential distribution; rather, for *k >* 1 in Eqs. 1 the eclipse stage duration is described by an Erlang distribution identified by its shape and scale, with a shape parameter greater than one. For illustrative purposes, in Fig. 6 we plot the distribution of the eclipse duration for *k* = 1 through to *k* = 10. One can see that the eclipse duration distribution for *k* = 2 resolves the major issue raised previously in modelling SHIV [35], namely, that with an exponential distribution, many cells instantly begin budding at the moment of virion attachment to the target cell. For *k >* 1, this issue is resolved as the probability of an eclipse duration of zero is negligible. We conclude that the simplest model that provides a statistically acceptable fit to the data has a shape parameter of at least *k* = 2, and find the strongest support for *k* = 3.

**Fig. 6.**
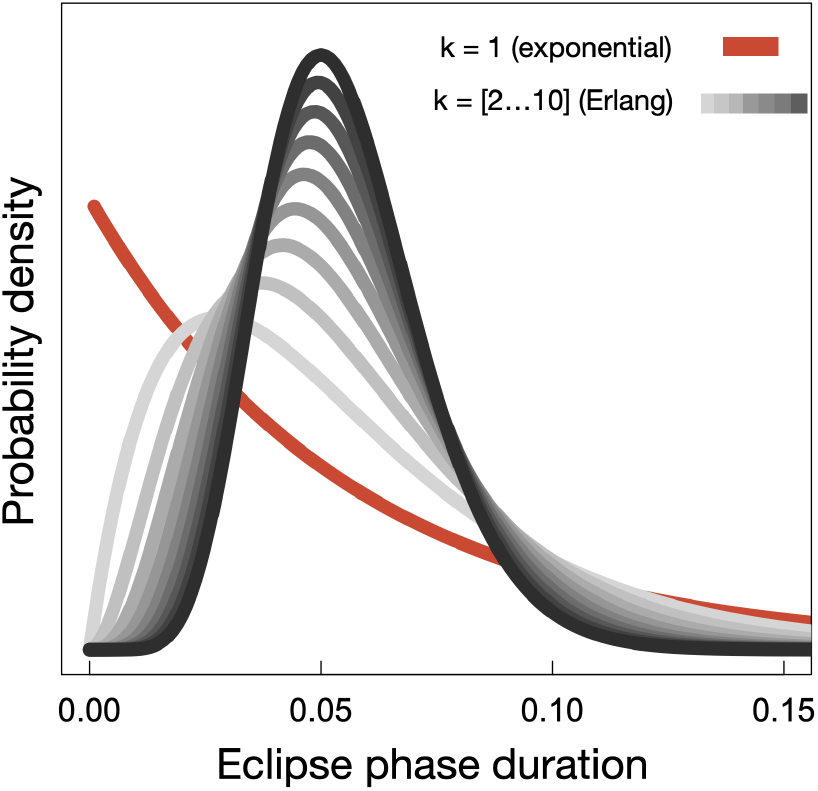
Eclipse duration distributions are shown for *k* = 1 (exponential, red line) and *k* = 2…10 (Erlang, grey lines). For *k* = 1, a substantial fraction of cells have near-zero eclipse times; this issue is resolved for *k >* 1.

Our finding that a target-cell limited model with *k >* 1 has more support compared to that with *k* = 1 mirrors findings for model fitting of other in-host infections [35, 55–57]. For example, recent rigorous modelling of the within-host kinetics of *Orthohantavirus* also concluded that *k >* 1 yielded more preferred models, where *k* = 3 was also found to have the strongest support [58]. Further, two-parameter distributions, such as Weibul, are considered more appropriate when stochastically modelling eclipse waiting times, where eclipse waiting times have been shown to have a significant affect on viral co-infection [59].

Despite these strong arguments for using *k >* 1 eclipse stages, we also analytically explored how varying *k* alters *R*_0_. We found that, as the number of eclipse stages *k* increased, *R*_0_ converged to 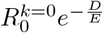. We also found that, for our model parameter values for *D* and *E* corresponding to the Jones, Néant, and Goyal data sets, the maximum decrease in *R*_0_ as *k* → ∞ is ∼ 5-8%. Thus previous studies of SARS-CoV-2 that assume a single eclipse compartment (*k* = 1) may see little impact on estimated *R*_0_ values if they were to instead use *k >* 1 eclipse stages (assuming the values of other fitted parameters remain constant). Indeed, where we consider fitting models consisting of one to ten eclipse stages, for our Jones fits (where *n* = 4344), we find very little variability in *R*_0_ across all *k* eclipse stages (Fig. 4a), and with the exception to *k* = 1, we find little variability with *k* in *R*_0_ estimated for the Néant data (*n* = 655) as well. We discuss the highly variable *R*_0_ estimated for the Goyal (*n* = 25) data set in more detail below.

We can compare our parameter results with those previously estimated for SARS-CoV-2. Previous in-host modelling work on SARS-CoV-2 viral load dynamics have utilized models with *k* = 1 eclipse stages [13, 15, 16, 28–30]. In this work, for our *k* = 1 model fits (Eqs. 1) we find good agreement in our fitted parameters with those previously reported for SARS-CoV-2. Our *k* = 1 parameter estimates for the Néant data set [13] agree well with those published by Néant *et al*. [13]. Furthermore, where Néant *et al*. [13] fix the clearance rate to *C* = 10, we fit this parameter and find *C* = 17.8 ± 7 *d*^*−*1^. We find a lower budding rate, B; where Néant *et al*. [13] find *B* = 6.08 × 10^5^ we find *B* = 4.88 × 10^4^. This discrepancy in *B* is most likely due to the differences in the time of infection, *t*_inf_, which we estimate to be -11.3 days versus the -4.8 days determined by Néant. For their target-cell limited model. Néant *et al*. [13] also find a mean of *R*_0_ = 36, where we find *R*_0_ = 41 for *k* = 1; however, across accepted models we find a much lower mean *R*_0_ of 9.4 (Fig. 4a).

The Néant data set consists of 655 SARS-CoV-2 positive hospitalized individuals with samples acquired between February and April of 2020 in France. The Jones data set consists of 4344 individuals from Berlin, 80% hospitalized, with samples acquired over a much longer time frame from February 2020 to April 2021. Thus, the Jones data set, consisting of more individuals, will not only have captured greater heterogeneity in SARS-CoV-2 in-host viral load dynamics (such as clearance and peak dynamics) but will also reflect a myriad of variants that swept through Germany over the course of 2020-2021. While the mean *R*_0_ we estimate using the Néant data set was 9.4, we find the mean *R*_0_ for the Jones data set to be 17.1. Referring to the best-ft parameter estimates in Fig. 3, we see that the higher *R*_0_ computed for the Jones fits is driven in part by a higher estimate of *α* for this data set. This higher attachment rate might be related to the increased infectivity of variants that emerged later in 2021 [60, 61].

We were also interested in exploring the reliability of model parameter estimation from low resolution data, where initial and peak dynamics may not have been captured. Consider the Goyal data set, which includes 25 individuals; times in these data correspond to days after a first positive test. It is known that symptom onset is correlated with peak infectious load where for SARS-CoV-2 symptoms have been shown to begin within 1 day of peak viral load [62]. Typically individuals will not seek testing until after symptoms have begun, thus, the Goyal data set likely captures late-stage dynamics, past the peak of infection; this conjecture is supported by the fact that the maximum viral load in this data set is considerably lower than that observed in the other data set. This limited data resolution poses an issue when trying to estimate parameters, such as the budding, clearance and per-target cell attachment rate, which are particularly important during the early-stage dynamics, as well as the initial time of infection. To illustrate the importance of sampling timescales on parameter estimation, we varied parameters for model Eqs. 1 around the mean values estimated for [27] to show their relative influence over the viral load dynamics (see Fig. 7). We observe that parameters *B* and *C* have a very similar influence over the slope of the initial rise in viral load. Similarly, *E* and *α* both influence the time to peak. Finally, we note that the post-peak slope remains at the same slope when any of these four parameters are changed; only *D* changes the slope of viral load decay. These conclusions are mirrored in the PRCC analysis (Fig.S5), which demonstrates that *B, C* and *E* strongly affect the peak viral load, while parameter *D* influences the peak density of budding cells.

**Fig. 7.**
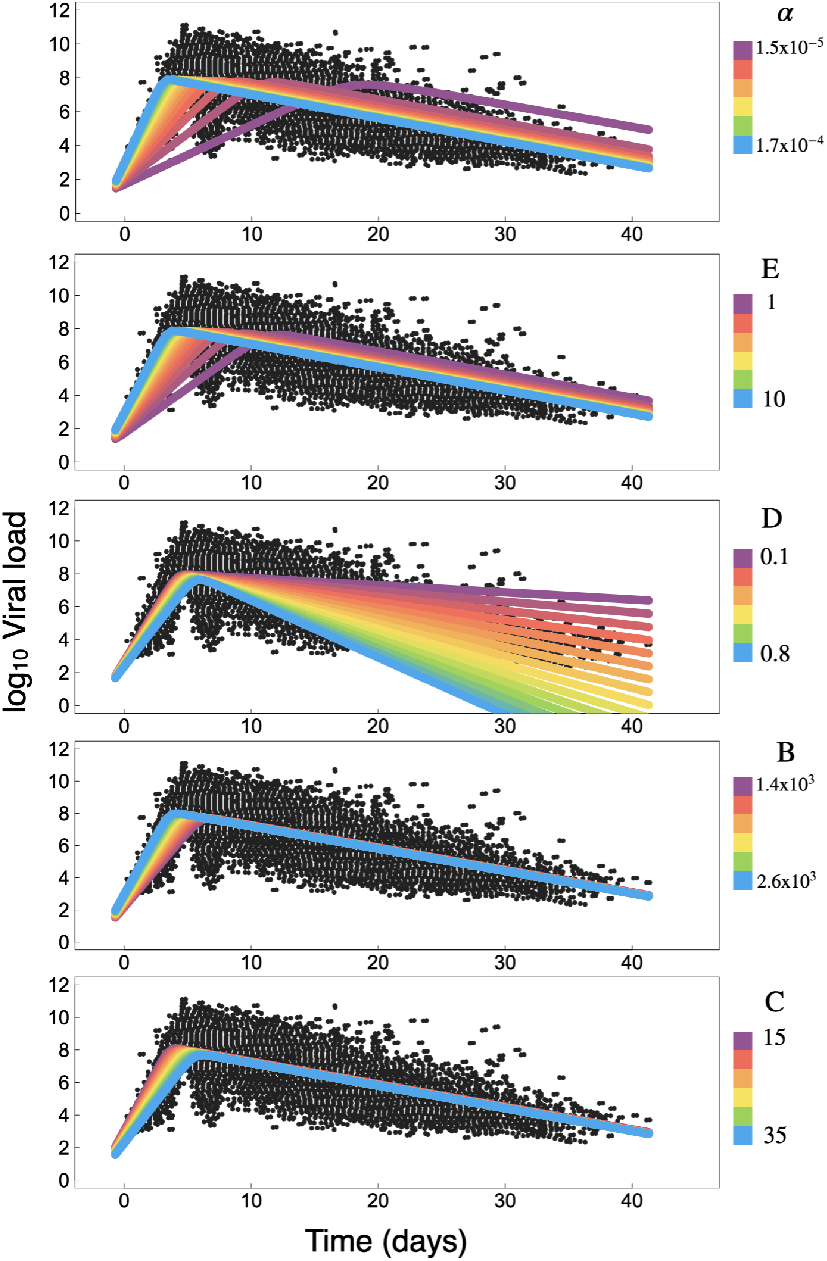
For illustrative purposes, parameters for model Eqs. 1 were varied around the mean values obtained for [27] to show their relative influence over the viral load model dynamics. We can see, for example, that the time-to-peak is largely influenced by *α* and *E*, the dynamics of the rise to the peak are influenced by *α, E, B* and *C*, while the slope of the viral load from peak to complete clearance is determined by *D*.

If initial infection dynamics are not present, it is therefore more difficult to estimate parameters without also considering parameter bounds and literature-informed priors. For data sets that do not capture the peak viral load, valid estimates of the parameter *D* should be possible, but the other parameters may not be uniquely identifiable. Taken together, Fig. 7 and Fig. 3 suggest that the parameter *C* is particularly hard to identify in these data, and cannot be quantified with precision; this is presumably because viral clearance is very fast when compared with the timescale of infected cell death. These parameter variations underlie the considerable variation in *R*_0_ values estimated for the Goyal data set, ranging from 18 to 36.5 as *k* changes (see Fig. 4). In a similar in-host study comprised of 13 SARS-CoV-2 positive individuals, Goncalves *et al*. determine an *R*_0_ of 8.6, but also report a large range of values of 1.9- 17.6 [28]. We propose that the large range of *R*_0_ values may be due to the absence of peak viral load data.

Are the parameters of our model uniquely identifiable in principle, given enough data? To address this question, we non-dimensionalize Eqs. 1 (Eqs. S16). We find that only three of the four values *EB, C, D*, and *E* are independently identifiable. However, in order to fit the reduced system of equations to data, both the time scaling (which involves *C*) and the viral load scaling (which involves *α*) are necessary. Thus, all the parameters are, in principle, meaningful to the data fitting performed here. To improve precision given limited data, one approach is to place bounds, or simply fix, particular parameters based on the relevant literature. Here, we limited *t*_inf_ to a maximum of 14 days, similar to Néant *et al*. [13], and also fixed the mean eclipse duration. Placing bounds on *t*_inf_ implicitly places bounds on the parameters that govern the rise to the peak, such as *B* and *C*.

While in-host modelling studies typically report residual errors (sensitivity of parameter estimates to measurement error), here we also carefully investigated the sensitivity of parameter estimates to model structure (Fig. 3). We also examined the sensitivity of predicted time courses (Fig. 7), the compound parameter *R*_0_ (Fig. 5), the standard error between model results and data (Fig. S2), and peak densities (Fig. S3) to variations in parameter estimates. These sensitivity analyses form a critical context for the interpretation of SARS-CoV-2 within-host parameter estimates, and should help to inform future modelling as more data become available.

## Data Availability

All data that support the findings of this study are available within the manuscript and the supplementary information files or from the corresponding authors upon reasonable request.

https://www.pnas.org/doi/suppl/10.1073/pnas.2017962118

http://www.ncbi.nlm.nih.gov/pubmed/34035154

## Acknowledgments

We thank Jeremie Guedj, and the Canadian In-host Modelling group (Dan Coombs, Morgan Craig, Thomas Hillen, Jude Kong, Kang Ling Liao, Nicole Mideo, Stephanie Portet, Angie Raad, James Watmough, and Samaneh Gholami) for fruitful discussions. This research is supported by NSERC Discovery Grants (JMH, LMW, IRM, MB), CIHR-Fields COVID Immunity Task Force (JMH), and an NRC Pandemic Response Challenge Program Grant (JMH, HKO). CSK acknowledges NSERC PGS-D funding.

## Declarations

We declare no competing interests.

## Notes

### Competing Interest Statement

The authors have declared no competing interest.

### Author Declarations

All data used in this manuscript has already been published (we are not publishing original data), and is openly available to the public. We used data obtained directly from their published source: [1]http://www.ncbi.nlm.nih.gov/pubmed/34035154 [2]https://www.science.org/doi/10.1126/sciadv.abc7112 [3]https://www.pnas.org/doi/suppl/10.1073/pnas.2017962118

