## Supplemental Material for "Multiple cohort study of hospitalized SARS-CoV-2 in-host infection dynamics: parameter estimates, sensitivity and the eclipse phase profile"

June 20, 2022

Chapin S. Korosec, Matt I. Betti, David W. Dick, Hsu Kiang Ooi, Iain R. Moyles, Lindi M. Wahl, Jane M. Heffernan

Corresponding author emails:

### Contents

**S1 Supporting Text** **2**

S1.1 Calculating $R_0$ . . . . . . 2

**S2 Parameter reduction and identifiability** **5**

**S3 Model parameter definitions, priors, and initial conditions** **7**

**S4 Parameter fit values** **8**

**S5 Time of Infection,** $t_{inf.}$ **9**

**S6 Sensitivity Analysis** **9**

### S1 Supporting Text

#### S1.1 Calculating $R_0$

We follow the method of Diekmann and Heesterbeek [1,2] to calculate $R_0$ for our target-cell limited model with $k$ eclipse stages,

$$\text{Target cells}: \frac{dy_T}{dt} = -\alpha y_T v \tag{S1a}$$

$$\text{Eclipse stage 1}: \frac{dy_1}{dt} = \alpha y_T v - (D + kE)y_1 \tag{S1b}$$

$$\text{Eclipse stage 2...k}: \frac{dy_j}{dt} = kEy_{j-1} - (D + kE)y_j, \quad j = 2...k \tag{S1c}$$

$$\text{Budding}: \frac{dy_B}{dt} = kEy_k - Dy_B \tag{S1d}$$

$$\text{Infectious Virions}: \frac{dv}{dt} = \epsilon By_B - \alpha y_T v - Cv \tag{S1e}$$

$$\text{Noninfectious Virions}: \frac{dw}{dt} = (1 - \epsilon)By_B - Cw. \tag{S1f}$$

$R_0$ is determined by considering a disease-free equilibrium scenario consisting of maximum susceptible cell population $y_{T_0}$ through computing the absolute value of the dominant eigenvalue of the next generation matrix, $\mathbf{G}$, where $\mathbf{G}$ is defined as the matrix product $\boldsymbol{T}\boldsymbol{\Sigma}^{-1}$. The Jacobian matrix $\boldsymbol{T}$ is the 'new-infection matrix' given by

$$\boldsymbol{T} = \frac{\partial T_i}{\partial x_j}(x_0). \tag{S2}$$

The Jacobian matrix $\Sigma$ is the disease transfer matrix, and represents the transfer of infected individuals between compartments. $\boldsymbol{\Sigma}$ is given by

$$\boldsymbol{\Sigma} = \frac{\partial \Sigma_i}{\partial x_j}(x_0). \tag{S3}$$

Here, $\boldsymbol{\Sigma}$ is more intuitively calculated by computing the sum $\boldsymbol{\Sigma} = \boldsymbol{\Sigma}^- - \boldsymbol{\Sigma}^+$, where $\boldsymbol{\Sigma}^-$ represents the transfer of infected cells out of each compartment, and $\boldsymbol{\Sigma}^+$ represents the transfer of infected cells into each compartment (neglecting the transfer of susceptible cells into the first eclipse stage, which is captured by $\boldsymbol{T}$). For the generalized case of $k$ eclipse stages, $\boldsymbol{T}$ and $\boldsymbol{\Sigma}$ are of size $(k+2\ x\ k+2)$. Here, we compute $R_0$ for $k = 5$.

The Jacobian matrices $\boldsymbol{T}$, $\boldsymbol{\Sigma}^+$ and $\boldsymbol{\Sigma}^-$ are

$$\boldsymbol{T} = \begin{bmatrix} 0 & 0 & 0 & 0 & 0 & 0 & \alpha y_{T_0} \\ 0 & 0 & 0 & 0 & 0 & 0 & 0 \\ 0 & 0 & 0 & 0 & 0 & 0 & 0 \\ 0 & 0 & 0 & 0 & 0 & 0 & 0 \\ 0 & 0 & 0 & 0 & 0 & 0 & 0 \\ 0 & 0 & 0 & 0 & 0 & 0 & 0 \\ 0 & 0 & 0 & 0 & 0 & 0 & 0 \end{bmatrix} \tag{S4}$$

$$\boldsymbol{\Sigma}^+ = \begin{bmatrix} 0 & 0 & 0 & 0 & 0 & 0 & 0 \\ KE & 0 & 0 & 0 & 0 & 0 & 0 \\ 0 & KE & 0 & 0 & 0 & 0 & 0 \\ 0 & 0 & KE & 0 & 0 & 0 & 0 \\ 0 & 0 & 0 & KE & 0 & 0 & 0 \\ 0 & 0 & 0 & 0 & KE & 0 & 0 \\ 0 & 0 & 0 & 0 & 0 & \epsilon B & 0 \end{bmatrix} \tag{S5}$$

$$\boldsymbol{\Sigma}^- = \begin{bmatrix} D+kE & 0 & 0 & 0 & 0 & 0 & 0 \\ 0 & D+kE & 0 & 0 & 0 & 0 & 0 \\ 0 & 0 & D+kE & 0 & 0 & 0 & 0 \\ 0 & 0 & 0 & D+kE & 0 & 0 & 0 \\ 0 & 0 & 0 & 0 & D+kE & 0 & 0 \\ 0 & 0 & 0 & 0 & 0 & D & 0 \\ 0 & 0 & 0 & 0 & 0 & 0 & \alpha y_{T_0} + C \end{bmatrix} \tag{S6}$$

Where $Q = \frac{\epsilon \alpha B y_{T_0}}{D(\alpha y_{T_0} + C)}$, the next-generation matrix is then determined to be

$$\boldsymbol{G} = \boldsymbol{T}[\boldsymbol{\Sigma}^{-} - \boldsymbol{\Sigma}^{+}]^{-1} = \begin{bmatrix} -\frac{Q(Ek)^5}{(D+kE)^5} & -\frac{Q(Ek)^4}{(D+kE)^4} & -\frac{Q(Ek)^3}{(D+kE)^3} & -\frac{Q(Ek)^2}{(D+kE)^2} & -\frac{Q(Ek)^1}{(D+kE)^1} & -Q & \frac{\alpha y_{T_0}}{\alpha y_{T_0}+C} \\ 0 & 0 & 0 & 0 & 0 & 0 & 0 \\ 0 & 0 & 0 & 0 & 0 & 0 & 0 \\ 0 & 0 & 0 & 0 & 0 & 0 & 0 \\ 0 & 0 & 0 & 0 & 0 & 0 & 0 \\ 0 & 0 & 0 & 0 & 0 & 0 & 0 \\ 0 & 0 & 0 & 0 & 0 & 0 & 0 \end{bmatrix}. \tag{S7}$$

The absolute value of the dominant (and in this case only non-zero) eigenvalue, $R_0$, is then

$$R_0 = \frac{\epsilon \alpha B y_{T_0} (5E)^5}{D(\alpha y_{T_0} + C)(D + 5E)^5}, \tag{S8}$$

For $k$ eclipse phases $R_0$ is

$$R_0^{k>1} = \frac{\epsilon \alpha B y_{T_0} (kE)^k}{D(\alpha y_{T_0} + C)(D + kE)^k}. \tag{S9}$$

In this work, we also consider the case with no eclipse stages ($k = 0$). The model then reduces to

$$\text{Target cells}: \frac{dy_T}{dt} = -\alpha y_T v \tag{S10a}$$

$$\text{Budding}: \frac{dy_B}{dt} = \alpha y_T v - D y_B \tag{S10b}$$

$$\text{Infectious Virions}: \frac{dv}{dt} = \epsilon B y_B - \alpha y_T v - C v \tag{S10c}$$

$$\text{Noninfectious Virions}: \frac{dw}{dt} = (1 - \epsilon) B y_B - C w, \tag{S10d}$$

for which we find the Jacobian matrices $\boldsymbol{T}$, $\boldsymbol{\Sigma}^{+}$ and $\boldsymbol{\Sigma}^{-}$ to be

$$\boldsymbol{T} = \begin{bmatrix} 0 & \alpha y_{T_0} \\ 0 & 0 \end{bmatrix} \tag{S11}$$

$$\boldsymbol{\Sigma}^{+} = \begin{bmatrix} 0 & 0 \\ \epsilon B & 0 \end{bmatrix} \tag{S12}$$

$$\boldsymbol{\Sigma}^{-} = \begin{bmatrix} D & 0 \\ 0 & \alpha y_{T_0} + C. \end{bmatrix} \tag{S13}$$

The next generation matrix is then

$$\boldsymbol{G} = \begin{bmatrix} \frac{\alpha y_{T_0} \epsilon B}{D(\alpha y_{T_0}+C)} & \frac{\alpha y_{T_0}}{\alpha y_{T_0}+C} \\ 0 & 0 \end{bmatrix} \tag{S14}$$

which gives the dominant eigenvalue

$$R_0^{k=1} = \frac{\alpha y_{T_0} \epsilon B}{D(\alpha y_{T_0} + C)}. \tag{S15}$$

### S2 Parameter reduction and identifiability

For a given value of $k$, the in-host model given in the main text (repeated here for reference) has six parameters: $\alpha$, $B$, $C$, $D$, $E$ and $\epsilon$.

$$\text{Target cells}: \frac{dy_T}{dt} = -\alpha y_T v \tag{S16a}$$

$$\text{Eclipse stage 1}: \frac{dy_1}{dt} = \alpha y_T v - (D + kE)y_1 \tag{S16b}$$

$$\text{Eclipse stage 2...k}: \frac{dy_j}{dt} = kEy_{j-1} - (D + kE)y_j, \quad j = 2...k \tag{S16c}$$

$$\text{Budding}: \frac{dy_B}{dt} = kEy_k - Dy_B \tag{S16d}$$

$$\text{Infectious Virions}: \frac{dv}{dt} = \epsilon By_B - \alpha y_T v - Cv \tag{S16e}$$

$$\text{Noninfectious Virions}: \frac{dw}{dt} = (1 - \epsilon)By_B - Cz_w. \tag{S16f}$$

By re-scaling each dependent variable and time, this model can be reduced to the following equivalent 4-parameter model:

$$\frac{dz_T}{d\tau} = -z_T z_v \tag{S17a}$$

$$\frac{dz_1}{d\tau} = z_T z_v - (\tilde{D} + k\tilde{E})z_1 \tag{S17b}$$

$$\frac{dz_j}{d\tau} = k\tilde{E}z_{j-1} - (\tilde{D} + k\tilde{E})z_j, \quad j = 2...k \tag{S17c}$$

$$\frac{dz_B}{d\tau} = z_k - \tilde{D}z_B \tag{S17d}$$

$$\frac{dz_v}{d\tau} = \epsilon\tilde{B}\tilde{E}z_B - z_T z_v - z_v \tag{S17e}$$

$$\frac{dz_w}{d\tau} = (1 - \epsilon)\tilde{B}\tilde{E}z_B - z_w, \tag{S17f}$$

where we let $\tau = Ct$, $z_T = \alpha y_T/C$, $z_i = \alpha y_i/C$ $(i = 1...k)$, $z_B = \alpha y_B/E$ and $z_v = \alpha v/C$. The composite parameters are $\tilde{D} = D/C$, $\tilde{E} = E/C$ and $\tilde{B} = B/C$.

System S17 is mathematically equivalent to system S16, and since the $z_w$ equation is decoupled, the dynamics of the system are determined by only three parameters, $\tilde{D}$, $\tilde{E}$ and $\epsilon\tilde{B}$. Thus only three of the four parameters $\epsilon B$, $C$, $D$ and $E$ are independently identifiable, in the sense that in the limit with infinite data available for populations $z_T$ through $z_v$ versus $\tau$, only the ratios $\epsilon B/C$, $D/C$ and $E/C$ could be determined. The parameter $\epsilon$ could be further estimated if data for $z_w$ were available.

However, in order to fit the reduced system to empirical data ($v + w$ versus $t$), both the time scaling (involving parameter $C$), and viral load scaling (involving parameter $\alpha$) are necessary. Thus ultimately all six parameters are informative in fitting system S16 to clinical datasets.

### S3 Model parameter definitions, priors, and initial conditions

All model assumptions, initial conditions, and definitions are listed in table S1.

| Parameter | Units | Definition | Prior | Random effects allowed? | Parameter Bounds | Comment |
|---|---|---|---|---|---|---|
| $\alpha$ | mL$\cdot d^{-1} \cdot$ copies$^{-1}$ | Per target cell attachment rate | $4.93x10^{-5}$ | Yes | No bounds | Prior informed by previous SARS-CoV-2 literature [3] |
| $D$ | $d^{-1}$ | Cell removal rate | 1 | Yes | No bounds | Fit. |
| $k$ | Unitless | Number of eclipse stages | N/A | No | N/A | Fixed to the number of eclipse stages |
| $E$ | $d^{-1}$ | Eclipse rate | 5 | No | N/A | Fixed for all fits. Value informed by [4] |
| $\epsilon$ | Unitless | fraction of infectious virions produced by budding cells | 0.001 | No | N/A | Fixed. Value referenced from [3, 5] |
| $B$ | copies$\cdot d^{-1} \cdot$ cell$^{-1}$ | Virion budding rate from infected cells | 8000 | Yes | No bounds | Prior informed by previous viral load literature [3] |
| $C$ | $d^{-1}$ | Virion clearance rate | 5 | Yes | No bounds | Fit. |
| $t_{inf.}$ | $d$ | Time of initial infection | 1 | Yes | 0.001-14 | Bounds on fit informed by previous SARS-CoV-2 literature [3] |
| | | | Initial Conditions (ICs) | | | |
| $y_{T0}$ | cells$\cdot$mL$^{-1}$ | Susceptible target cell IC | $1.33x10^5$ | No | N/A | Fixed. referenced from [3] |
| $y_{i0}$ | cells$\cdot$mL$^{-1}$ | ith eclipse state IC | 0 | No | N/A | Fixed |
| $y_{B0}$ | cells$\cdot$mL$^{-1}$ | Budding cells IC | $\frac{1}{30}$ | No | N/A | Fixed, referenced from [3] |
| $v_0$ | copies$\cdot$ml$^{-1}$ | Infectious virion IC | 0 | No | N/A | Fixed |
| $F_0$ | copies$\cdot$ml$^{-1}$ | Non-infectious virion IC | 0 | No | N/A | Fixed |

Table S1: Model parameters definitions, initial conditions, and prior assumptions for all model fits.

#### S4 Parameter fit values

| Data set [ref.] | Parameter | Eclipse Stage, $k$ | | | | | | | | | | | Mean | Standard Deviation |
|---|---|---|---|---|---|---|---|---|---|---|---|---|---|---|
| | | 0 | 1 | 2 | 3 | 4 | 5 | 6 | 7 | 8 | 9 | 10 | | |
| Goyal [6] | $\alpha$ | 5.0x10$^{-5}$ | 5.1x10$^{-5}$ | 1.7x10$^{-3}$ | 1.6x10$^{-3}$ | 5.0x10$^{-3}$ | 4.8x10$^{-3}$ | 2.4x10$^{-3}$ | 1.6x10$^{-3}$ | 7.5x10$^{-3}$ | 2.8x10$^{-3}$ | 6.6x10$^{-3}$ | 3.1x10$^{-3}$ | 2.5x10$^{-3}$ |
| | $\epsilon B$ | 31.8 | 76.0 | 13.8 | 17.1 | 9.8 | 10.5 | 16.0 | 17.5 | 10.3 | 18.6 | 10.3 | 21.1 | 19.3 |
| | $C$ | 19.95 | 39.95 | 27.1696 | 38.12 | 33.74 | 26.24 | 41.25 | 29.1011 | 41.15 | 40.78 | 45.55 | 34.8 | 8.1 |
| | $D$ | 0.52 | 0.55 | 0.47 | 0.46 | 0.46 | 0.48 | 0.46 | 0.48 | 0.46 | 0.46 | 0.45 | 0.48 | 0.03 |
| | $t_{inf.}$ | -13.93 | -14.0 | -13.96 | -13.93 | -13.94 | -13.95 | -13.94 | -13.95 | -13.88 | -13.85 | -13.91 | -13.90 | 0.04 |
| Jones [7] | $\alpha$ | 4.9x10$^{-5}$ | 5.1x10$^{-5}$ | 9.01x10$-$5 | 7.9x10$^{-5}$ | 1.6x10$^{-4}$ | 1.7x10$^{-4}$ | 7.8x10-5 | 8.25x10$^{-5}$ | 4.8x10$^{-4}$ | 8.1x10$^{-5}$ | 1.7x10$^{-4}$ | 1.4x10$^{-4}$ | 1.2x10$^{-4}$ |
| | $\epsilon B$ | 13.2 | 32.0 | 14.4 | 19.5 | 8.7 | 8.7 | 21.1 | 19.5 | 6.3 | 20.1 | 8.9 | 15.7 | 7.6 |
| | $C$ | 18.5 | 34.2 | 18.4 | 25.1 | 11.0 | 10.9 | 26.8 | 25.2 | 7.9 | 25.8 | 11.2 | 19.5 | 8.5 |
| | $D$ | 0.32 | 0.33 | 0.32 | 0.32 | 0.32 | 0.32 | 0.32 | 0.32 | 0.32 | 0.32 | 0.32 | 0.321 | 0.003 |
| | $t_{inf.}$ | -1.05 | -1.12 | -1.20 | -1.19 | -1.25 | -1.24 | -1.19 | -1.21 | -1.27 | -1.2 | -1.26 | -1.20 | 0.06 |
| Neant [3] | $\alpha$ | 4.9x10$^{-5}$ | 4.8x10$^{-5}$ | 4.0x10$^{-5}$ | 4.5x10$^{-5}$ | 5.2x10$^{-5}$ | 5.0x10$^{-5}$ | 4.2x10$^{-5}$ | 4.1x10$^{-5}$ | 6.1x10$^{-5}$ | 5.3x10$^{-5}$ | 5.4x10$^{-5}$ | 4.9x10$^{-5}$ | 6.3x10$^{-6}$ |
| | $\epsilon B$ | 21.3 | 48.8 | 14.4 | 18.6 | 9.5 | 10.1 | 20.7 | 18.3 | 10.1 | 19.7 | 10.3 | 18.3 | 11.1 |
| | $C$ | 18.7 | 17.8 | 23.3 | 32.4 | 17.4 | 17.7 | 35.6 | 28.6 | 20.4 | 33.4 | 19.8 | 24.1 | 7.0 |
| | $D$ | 0.29 | 0.30 | 0.30 | 0.29 | 0.29 | 0.29 | 0.29 | 0.29 | 0.29 | 0.30 | 0.29 | 0.292 | 0.004 |
| | $t_{inf.}$ | -9.37 | -11.31 | -13.60 | -12.65 | -13.28 | -13.29 | -13.09 | -13.16 | -12.34 | -11.59 | -13.03 | -12.4 | 1.2 |

Table S2: All fitted parameter values for Goyal, Jones, and Neant cohorts for all eclipse stages, $k$, examined in this work. Associated errors on each parameters are shown on Figure 2. The mean and standard deviation listed at computed across all respective parameters shown for all $k$ values.

#### S5 Time of Infection, $t_{inf.}$

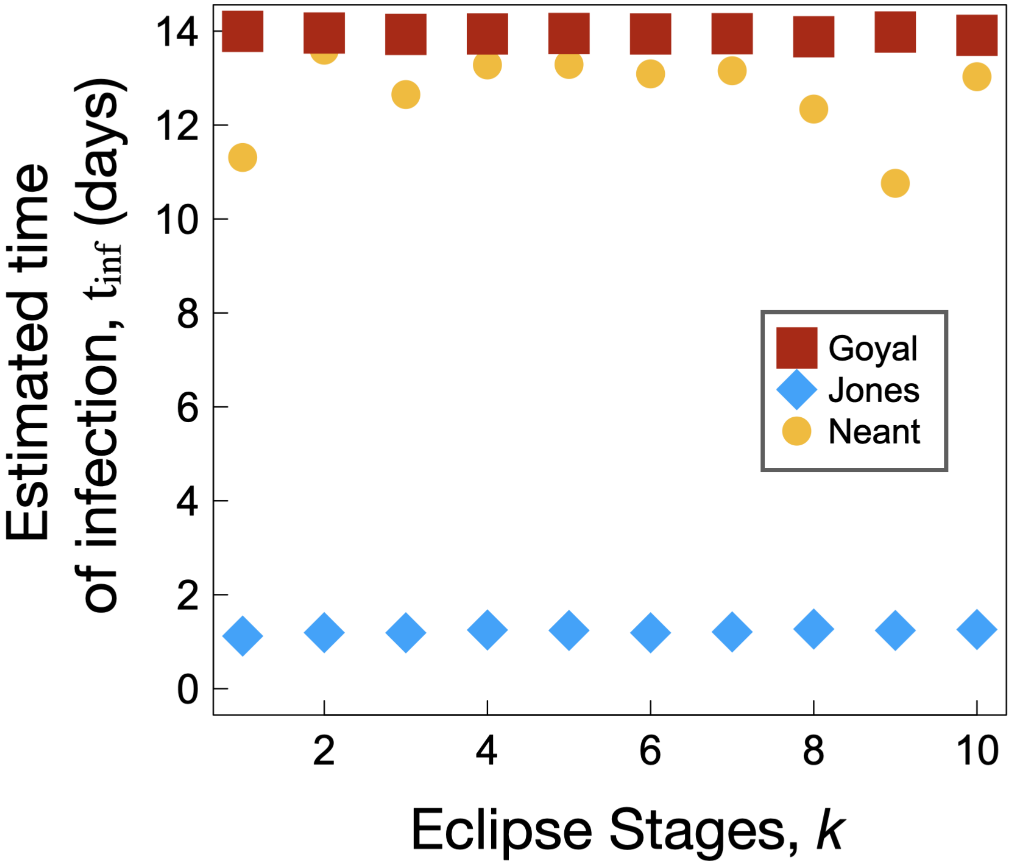


Figure S1: Estimated time of infection, $t_{inf.}$, for the Neant, Goyal, and Jones data sets for all $k$ eclipse stages.

#### S6 Sensitivity Analysis

To complement the error analysis and log-likelihood fit criteria produced by Monolix, we complete an independent error analysis on the fitted parameters $\alpha$, $B$, $C$, and $D$ and PRCC sensitivity analysis on the model parameters $\alpha$, $B$, $C$, $D$, and $E$ (See methods for details).

Fig. S2a shows an example output of the mean standard error as a function of $\epsilon B$, where $\epsilon B$ is varied uniformly from 1 to 50. The Monolix fit value for the $k = 5$ Jones data set is shown as a red solid line (which we denote by $B_M$), where the blue region flanked by red dashed lines represents the area captured by the Monolix error analysis ($B_M - \sigma_{B_M}$ to $B_M + \sigma_{B_M}$). The fitted value is close to a minimum in standard error, where the actual minimum is captured within the blue region. Fig. S2b displays boxplots of all standard errors computed for each respective eclipse stage. Here we vary each fitted parameter from its Monolix-reported minimum to maximum value according to the reported parameter error from the fits, $\sigma$. The parameter combinations resulting in the largest mean standard error, for all $k$, correspond to the minimum $\alpha$ and $B$ of $\alpha_M - \sigma_{\alpha_M}$, and $B = B_M - \sigma_{B_M}$, respectively, and maximum $D$ and $C$ of $D = D_M + \sigma_{D_M}$, and $C = C_M + \sigma_{C_M}$, respectively. The parameter combinations resulting in the minimum mean standard error, for all $k$, correspond to the maximum $\alpha$ and $B$, of $\alpha = \alpha_M + \sigma_{\alpha_M}$, $B = B_M + \sigma_{B_M}$, respectively, while $C$ and $D$ are the Monolix-reported values, $D = D_M$, and $C = C_M$, respectively.

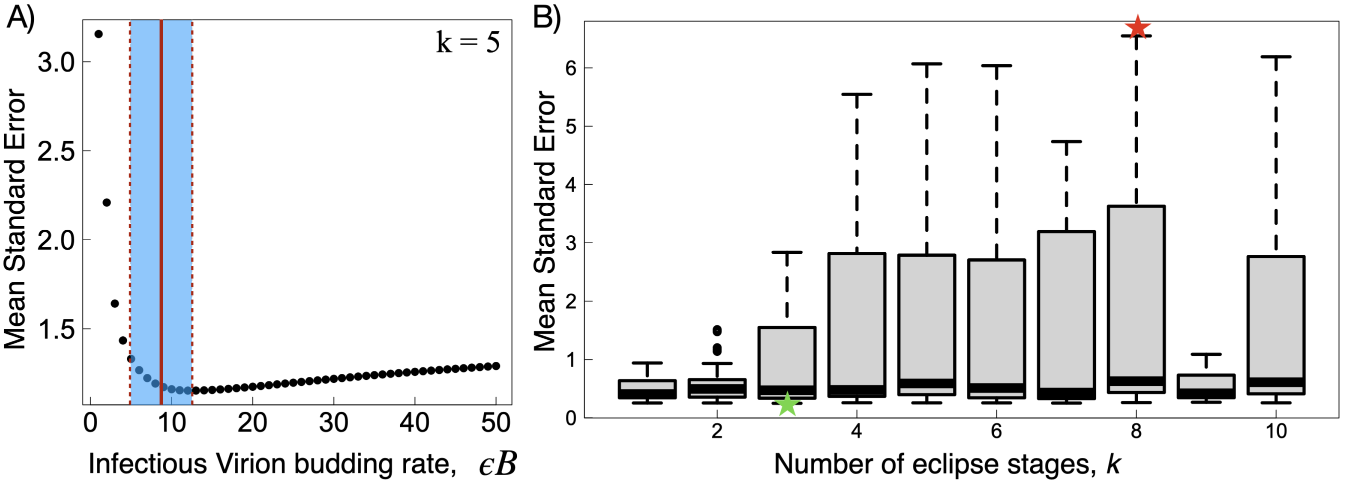


Figure S2: Standard error analysis for fitted parameters. (A). Mean standard error as a function of virion budding rate, $\epsilon B$, for $k = 5$, Jones data set. Red solid line is the Monolix-fit-determined value, while the blue region encompasses the error on $B$ from the fit. (B) Boxplot analysis displaying the spread in mean standard error for all parameter combinations for all eclipse stages. The red star labels the maximum error, corresponding to $k = 8$, $\alpha = \alpha_M - \sigma_{\alpha_M}$, $B = B_M - \sigma_{B_M}$, $D = D_M + \sigma_{D_M}$, and $C = C_M + \sigma_{C_M}$. The green star labels the minimum error, corresponding to $k = 3$, $\alpha = \alpha_M + \sigma_{\alpha_M}$, $B = B_M + \sigma_{B_M}$, $D = D_M$, and $C = C_M$.

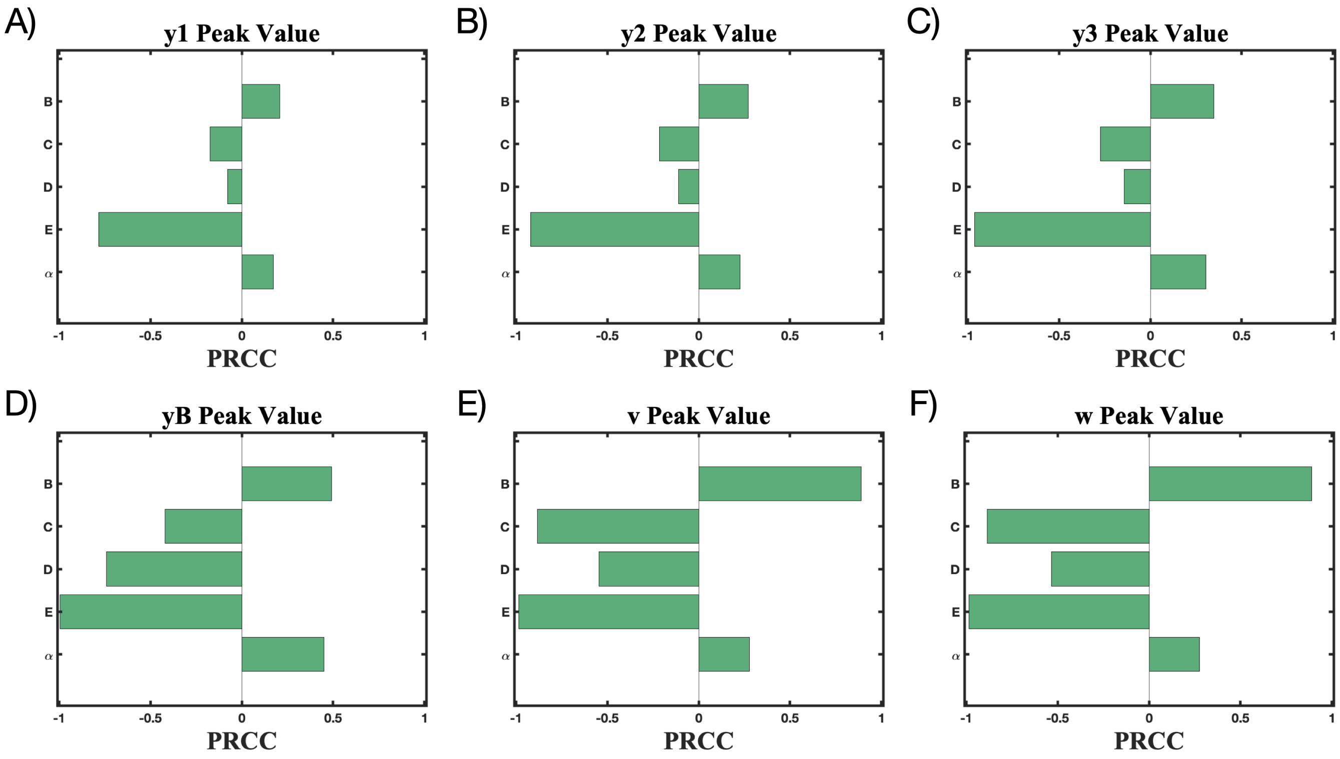


Figure S3: PRCC analysis was employed to study the effects of model outcomes on the peak value of each state variable. Fitted parameters $B$, $C$, $D$ and $\alpha$ are examines, additionally $E$ is examined although $E$ was fixed for all fits in this work. Model Eqs. S1 are used with $k = 3$ eclipse stages. A-F) display the results of the PRCC analysis for $y_1$, $y_2$, $y_3$, $y_B$, $v$, and $w$, respectively.
